# Suppression of de novo antibody responses against SARS-CoV2 and the Omicron variant after mRNA vaccination and booster in patients with B cell malignancies undergoing active treatment, but maintenance of pre-existing antibody levels against endemic viruses

**DOI:** 10.1101/2022.03.17.22272389

**Authors:** Joseph Azar, John P. Evans, Madison Sikorski, Karthik Chakravarthy, Selah McKenney, Ian Carmody, Cong Zeng, Rachael Teodorescu, No Joon Song, Jamie Hamon, Donna Bucci, Maria Velegraki, Chelsea Bolyard, Kevin P. Weller, Sarah Reisinger, Seema A. Bhat, Kami J. Maddocks, Richard J. Gumina, Anastasia N. Vlasova, Eugene M. Oltz, Linda J. Saif, Dongjun Chung, Jennifer A. Woyach, Peter G. Shields, Shan-Lu Liu, Zihai Li, Mark P. Rubinstein

**Author notes:** Co-corresponding authors Corresponding authors: Jennifer A. Woyach, Peter G. Shields, Shan-Lu Liu, Zihai Li, Mark P. Rubinstein. These authors contributed equally (Joseph Azar, John P. Evans, Madison Sikorski, Karthik Chakravarthy).

## Abstract

The impact of SARS-CoV2 vaccination in cancer patients remains incompletely understood given the heterogeneity of cancer and cancer therapies. We assessed vaccine-induced antibody response to the SARS-CoV2 Omicron (B.1.1.529) variant in 57 patients with B cell malignancies with and without active B cell-targeted therapy. Ancestral- and Omicron-reactive antibody levels were determined by ELISA and neutralization assays. In over one third of vaccinated patients at the pre-booster timepoint, there were no ELISA-detectable antibodies against either the ancestral strain or Omicron variant. The lack of vaccine-induced antibodies was predominantly in patients receiving active therapy such as anti-CD20 monoclonal antibody (mAb) or Bruton’s tyrosine kinase inhibitors (BTKi). While booster immunization was able to induce detectable antibodies in a small fraction of seronegative patients, the benefit was disproportionately evident in patients not on active therapy. Importantly, in patients with post-booster ELISA-detectable antibodies, there was a positive correlation of antibody levels against the ancestral strain and Omicron variant. Booster immunization increased overall antibody levels, including neutralizing antibody titers against the ancestral strain and Omicron variant; however, predominantly in patients without active therapy. Furthermore, ancestral strain neutralizing antibody titers were about 5-fold higher in comparison with those to Omicron, suggesting that even with booster administration, there may be reduced protection against the Omicron variant. Interestingly, in almost all patients regardless of active therapy, including those unable to generate detectable antibodies against SARS-CoV2 spike, we observed comparable levels of EBV, influenza, and common cold coronavirus reactive antibodies demonstrating that B cell-targeting therapies primarily impair de novo but not pre-existing antibody levels. These findings suggest that patients with B cell malignancies on active therapy may be at disproportionately higher risk to new versus endemic viral infection and suggest utility for vaccination prior to B cell-targeted therapy.

## Introduction

The latest SARS-CoV2 variant, Omicron (B.1.1.529), exhibits over 30 mutations in the Spike protein, and 15 mutations in the receptor binding domain (RBD), relative to the ancestral strain, (1-3). These mutations may allow evasion of SARS-CoV2 immunity, particularly antibody responses, and therefore, overcome vaccine-mediated immunity in some individuals (2, 4, 5). While emerging data suggest that in healthy immunized individuals there is sufficient immune protection against serious disease (6), the level of vaccine-induced protection in immuno-compromised individuals such as cancer patients is not well known. This is of particular importance to patients with hematological malignancies who have a higher risk of death from COVID-19 than solid tumor patients (7, 8), and for whom emerging data suggest continued elevated risk from COVID-19 even after vaccination (9, 10). A third dose of the mRNA vaccine (booster) can augment antibody responses in cancer and organ transplant patients who previously received a primary (2-dose) mRNA vaccine (11-14). Furthermore, we and others have reported that in healthy individuals and solid tumor patients, booster immunization can substantively augment neutralizing antibodies including against the Omicron variant (2-4, 11, 15-19). However, comparatively little is known about the impact of booster immunization in highly immune-compromised patients such as those with B cell malignancies, particularly those patients on active therapy. Such immune impairment may be particularly problematic for chronic lymphocytic leukemia (CLL) and non-Hodgkin’s lymphoma (NHL) patients, in whom B cell-targeted therapies have been shown to impair vaccine-mediated antibody responses (12, 17, 20-24). While recent studies suggest that booster immunization is less effective in patients with hematological malignancies, including the induction of Omicron neutralizing antibodies (12, 17, 23, 25), these studies provided only a limited assessment of the impact of booster immunization on antibody levels in patients with or without active B cell-targeted therapy. Here we evaluate the impact of booster immunization on levels of antibodies specific for Omicron and ancestral variants in cancer patients with or without active B cell-targeted therapy. Furthermore, we assess the extent to which B cell-targeted therapies impair de novo versus pre-existing antibody-mediated immunity.

## Results

### Patients and therapies received

Our goal was to assess mRNA vaccine (mRNA-1273 or BNT162b2)-induced antibody responses to Omicron (B.1.1.529) in comparison with ancestral SARS-CoV2 in a highly immune compromised patient population. Accordingly, we evaluated 57 patients with CLL (n=35) and NHL (n=22) who had paired serum samples before and after booster immunization (**Table 1**). Thirty of the 57 patients were on active therapy including 6 patients on anti-CD20 or anti-BAFF mAb, 15 patients on Bruton tyrosine kinase (BTK) inhibitors, and 7 patients receiving the combination of these agents. Six of the thirty patients stopped active therapy before booster immunization with drug stoppage being in a range of 4 months before to 4 months after primary vaccination. Of the remaining 27 patients without active therapy, 9 were untreated for their disease and the remaining 18 had received prior B cell-targeting therapies (mean = 4.9 years, range 15 months to 15 years). The first paired sample was collected 66-216 days (mean 139) after the primary vaccination (2^nd^ mRNA vaccine dose) and the second paired sample was collected 5-112 days (mean 52) after the booster vaccine. The time range between first and second sample collection was 21-182 days (mean 94). Patient demographic details are described in **Table 1**.

**Table 1.**
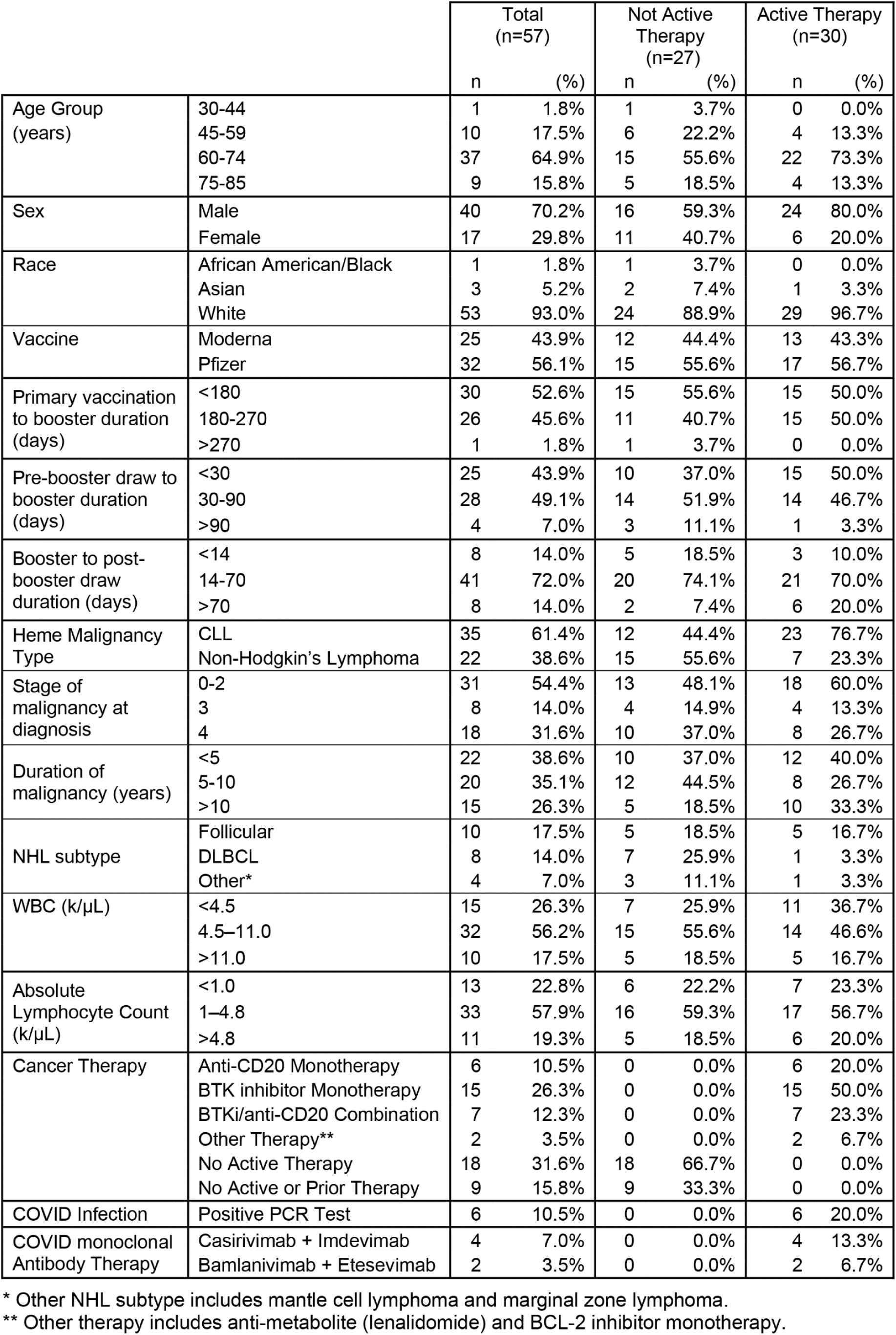

### Compromised antibody levels in boosted cancer patients undergoing B cell-targeted therapy

As a first step in understanding how targeted B cell therapies impact SARS-CoV2 humoral responses following vaccination and/or boosting, we determined the proportion of patients who were seronegative for antibodies ancestral and Omicron spike protein using an enzyme linked immunosorbent assay (ELISA). After primary immunization but before booster administration, we found the proportion of patients seronegative for both ancestral and Omicron spike was significantly elevated in those who received active therapy versus those not receiving active therapy (53% (16 of 30 patients) versus 18% (5 of 27 patients), p<0.05) (**Fig. 1**). Upon booster immunization, the lack of detectable antibodies continued to be disproportionate in patients with active therapy versus those not receiving active therapy (40% (12 of 30 patients) versus 7.4% (2 of 27), p<0.01) (**Fig. 1A**). Similar results were obtained using ELISA to detect antibodies reactive against ancestral and Omicron RBD (**Fig. 1B**). Of patients who were seronegative for either ancestral or Omicron spike before booster administration, only 33% (7 of 21) converted to seropositivity post-booster administration. Similarly, 19% (5 of 26) of patients without detectable antibodies against either ancestral or Omicron RBD seroconverted post-booster administration. Of the 6 patients that stopped active therapy prior to booster administration, all were seronegative before and after booster administration, and all had been treated with anti-CD20 or anti-BAFF mAb. Importantly, upon analysis of patients with detectable antibodies against spike or RBD, there was a positive correlation between antibody levels against the ancestral and Omicron spike and RBD independent of treatment (**Fig. 2**). For example, there was a positive correlation after booster administration between ancestral RBD and Omicron RBD in patients with (r=0.83) or without (r=0.82) active therapy. Collectively, these results demonstrate a severe deficiency in the ability to develop vaccine-associated antibodies in cancer patients with active B cell-targeted therapies, but highlight that patients with detectable antibodies exhibit substantial overlap in reactivity between the ancestral and Omicron SARS-CoV2 variant.

**Figure 1:**
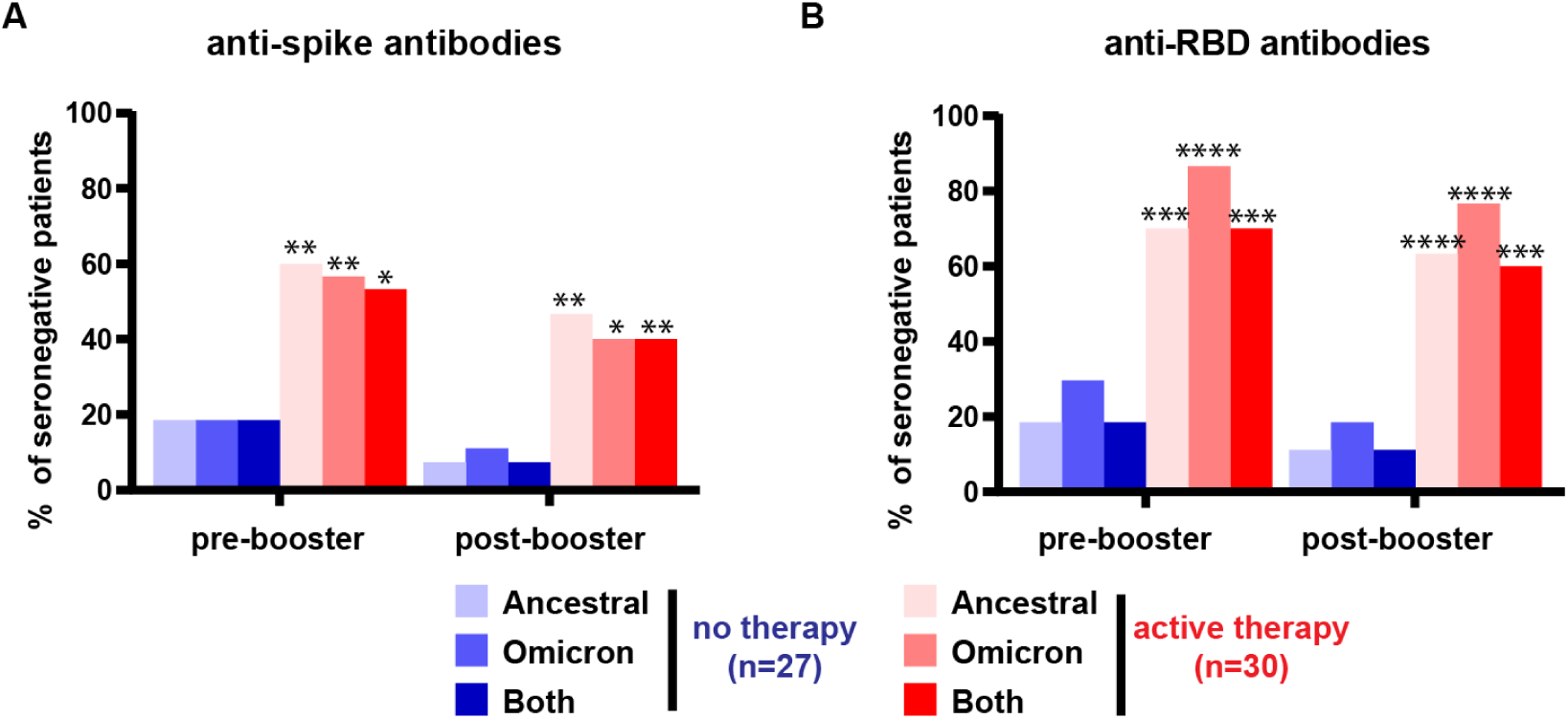
Frequency of patients without antibodies to the ancestral, Omicron, or both for SARS-CoV2 spike and RBD protein. (A) Each bar depicts the percentage of patients deficient in detectable antibodies against the ancestral RBD, Omicron RBD, or antibodies recognizing either target as determined by ELISA. (B) Same as ‘A’, except antibodies against ancestral spike, Omicron spike, or antibodies recognizing either target. Fisher’s exact test was conducted to compare the number of seronegative patients with no active therapy and active therapy. Shown are comparisons between active therapy and no therapy for the same antibody evaluation and group, as indicated by the arrow. *, **, *** and **** to indicate p< 0.05, 0.01, 0.001, 0.0001. Fisher’s exact test was also conducted to compare ancestral, omicron, or both within individual groups, and the differences were not statistically significant.

**Figure 2:**
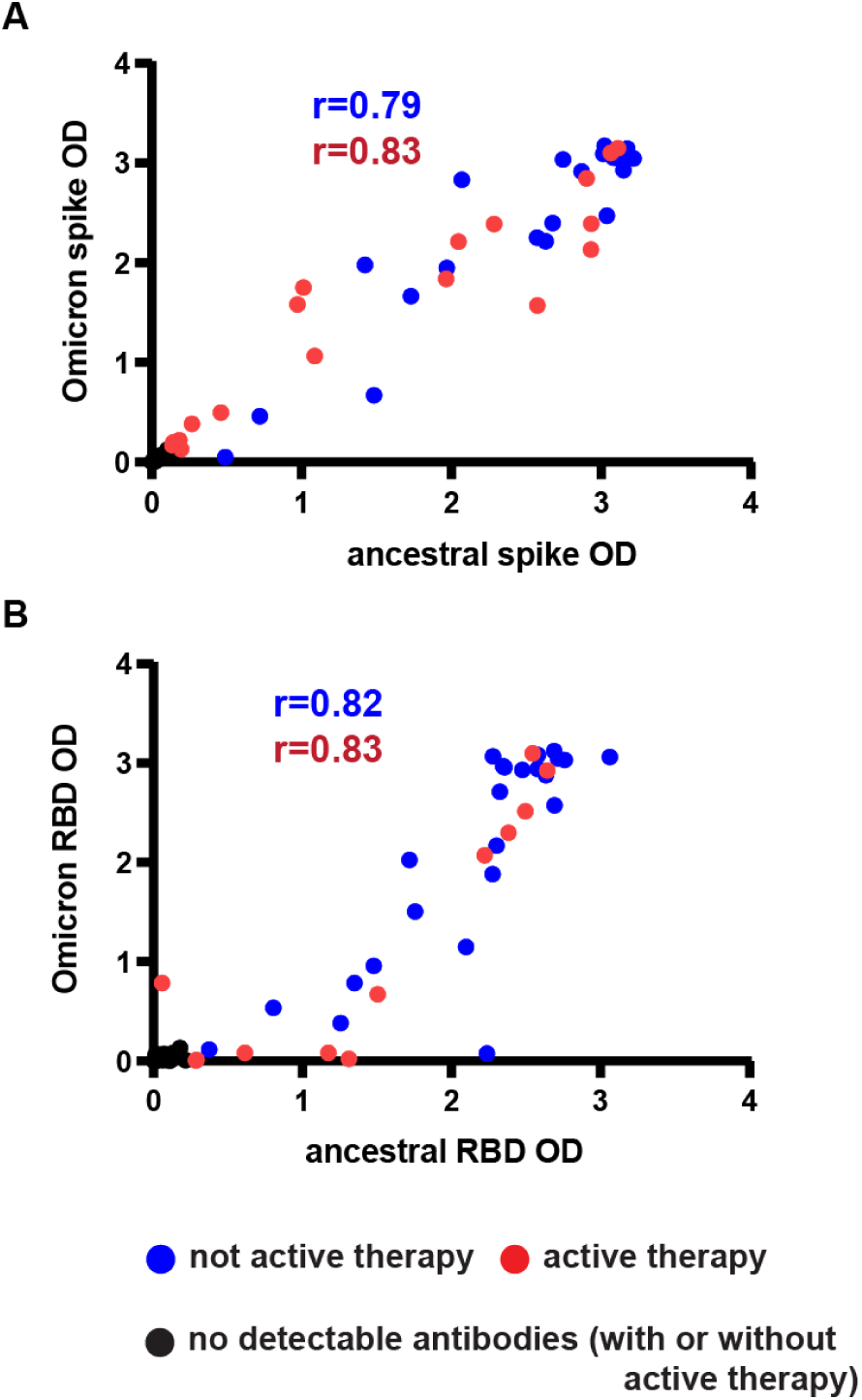
Positive correlation between antibody levels against ancestral and Omicron spike and RBD post-booster. (A) Dot plot shows the post-booster patient timepoints graphed by Omicron spike versus ancestral spike as determined by ELISA. The red dots depict patients on active therapy, the blue dots depict patients not on active therapy, and the black dots indicate patients without any detectable antibodies. (B) Same as ‘A’, except Omicron RBD versus ancestral RBD. The Spearman’s rank correlation coefficient was calculated for patients on active therapy (red) or not on active therapy (blue) with exclusion of patients without detectable antibodies..

We next assessed relative levels of antibodies against ancestral and Omicron spike across all patients (**Fig. 3A-C**). While booster immunization clearly enhanced both ancestral and Omicron-reactive spike antibody levels in patients without active therapy (3394 vs 5507, p<0.0001 for ancestral spike, and 2844 versus 5322, p<0.0001 for Omicron spike) (**Fig. 3B**), results for patients with active therapy were less clear-cut. Although there was a trend for increased levels of ancestral-reactive spike antibodies following booster administration in patients with active therapy, this trend was not statistically significant. However, for Omicron-reactive spike antibodies, the post-booster increase was significant (809 vs 1702, p<0.05) (**Fig. 3C**). Notably, for patients on active therapy, 23% (7 of 30) showed clear increases in antibodies against both ancestral and Omicron spike, suggesting a subset of patients benefitted from the booster. The impact of the booster was similar on ancestral- and Omicron-reactive RBD antibodies (**Fig. S1**). In addition to the deficiency in booster-mediated immunity apparent in patients on active therapy, these ELISA results confirm previous findings that patients with active therapy are deficient in generating antibodies after primary vaccination (12, 17, 20-24). Thus, pre-booster antibody levels in patients with active therapy were significantly reduced for all targets evaluated by ELISA, including ancestral spike (p<0.001), Omicron spike (p<0.001), ancestral RBD (p<0.001), and Omicron RBD (p<0.01) (**Fig. S1**). For an illustrative comparison of antibody responses between patients, we subdivided patients into 4 groups based on their antibody response to ancestral RBD (**Fig. S2A**). We observed, similar patterns of antibody levels pre- and post-booster against ancestral RBD, Omicron RBD, ancestral spike, and Omicron spike, again suggesting the ability of patients to develop cross-reactive antibodies. We also evaluated IgA and IgM antibody responses against the ancestral Spike protein (**Fig. S2B**). While we observed post-booster increased levels of spike-reactive IgA (p<0.001) and IgM (p<0.001) in individuals without active therapy, there was not a statistically significant increase in these antibodies for patients undergoing active treatment. Notably, IgG was the dominant antibody detected, and we did not observe patients with IgA or IgM in the absence of IgG (**Fig. S2B**).

**Figure 3:**
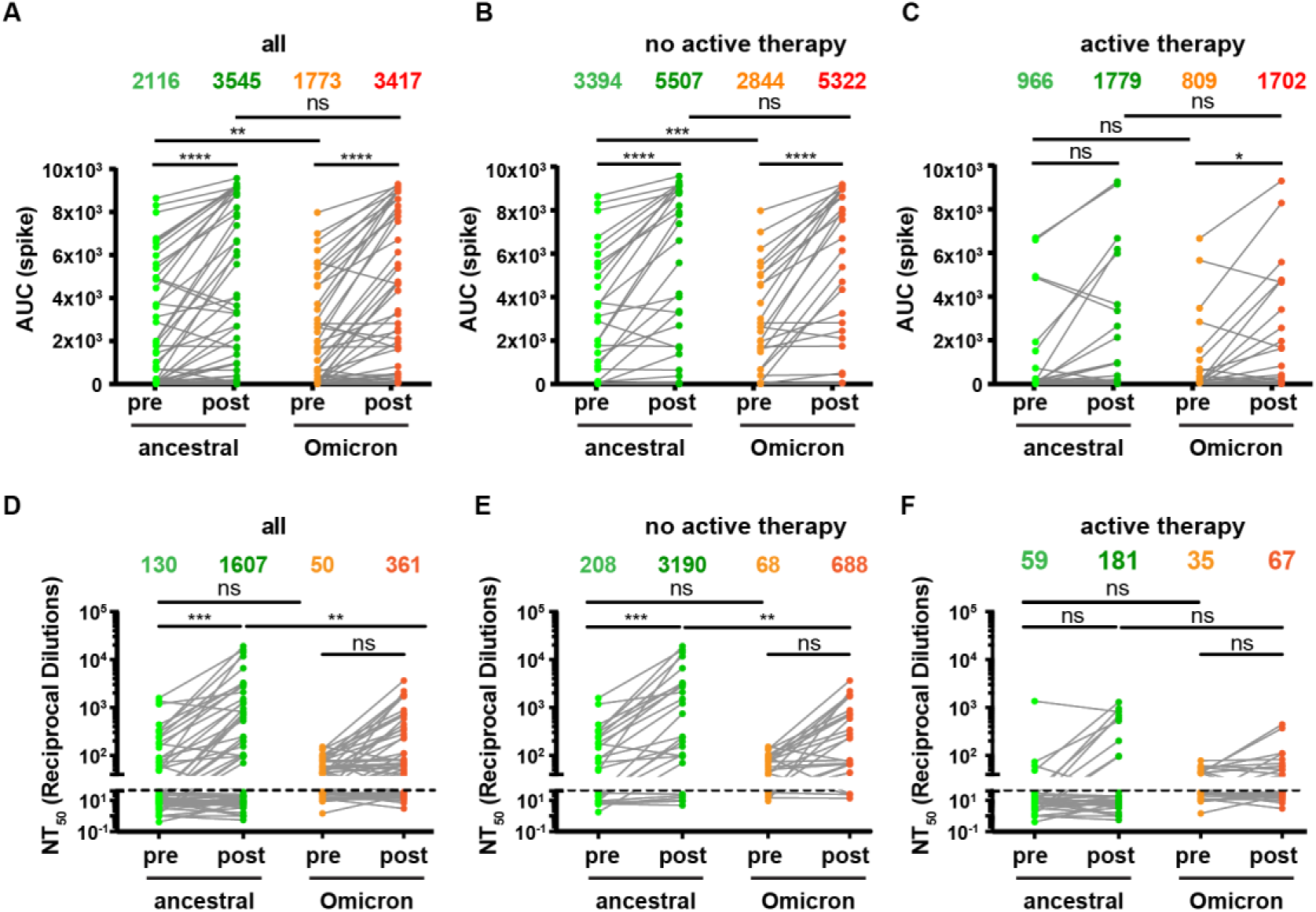
ELISA and viral neutralization assay depict variable antibody response to the ancestral and Omicron SARS-CoV2 strains before and after booster vaccination in hematological cancer patients. (A-C) Shows area under the curve (AUC) for all patients ‘A’, patients without active therapy ‘B’, and for patients with active therapy ‘C”. (D-F) The same as ‘A-C’, except showing the 50% neutralization titer (NT50). For all data, each symbol represents a timepoint that serum was isolated and the line connects individual patients. The number above each graph represent arithmetic mean. ANOVA was used to compare between pre-booster ancestral, post-booster ancestral, pre-booster Omicron, post-booster Omicron NT50/AUC values of the same patient *,**, ***, **** indicate p< 0.05, 0.01, .001, 0.0001.

### Booster-augmented neutralizing antibodies against the SARS-CoV2 ancestral and Omicron variant are increased in B cell malignancy patients without active therapy

To determine if antibody levels correlated with functional blockade of SARS-CoV2 pseudovirus, we assessed neutralizing antibody response to the ancestral and Omicron variant of SARS-CoV2 using our previously reported pseudotyped lentivirus neutralization assay (11, 26). In these experiments, we used ancestral D614G as a reference for comparison, and determined the change in 50% neutralization titer (NT_50_) before and after booster in all patients (**Fig. 3D**). For the ancestral strain, there was a 12-fold increase in NT_50_ (from 130 to 1607; p < 0.001) before and after booster immunization. In contrast, for the Omicron variant, there was only a 7-fold increase in NT_50_ (from 50 to 361); the latter increase was not significant, possibly due to the fact that 39% (22/57) of patients had an NT_50_ titer below the level of detection against Omicron following booster vaccine administration. Notably, NT_50_ titers against the Omicron strain post-booster exhibited a 4.5-fold reduction (1607 vs 361, p<0.001) compared to the ancestral strain consistent with previous findings (2-4, 11, 15-17).

In keeping with our ELISA results, patients not on active therapy had substantially higher titers of neutralizing antibodies after booster administration (**Fig. 3E and F**). Thus, for patients without active therapy, there was a 15-fold increase in NT_50_ (208 to 3190; p<0.001) in ancestral neutralizing antibodies (**Fig. 3E**). In comparison, after booster administration, there was a 11.5-fold increase in NT_50_ in Omicron neutralizing antibodies (68 to 688) although this difference did not attain statistical significance. In patients with active B cell-targeting therapies, there was a minimal increase in ancestral or Omicron neutralizing antibodies, and these differences were not significant. However, there appeared to be a small subset of active treatment patients who benefited from the booster (**Fig. 3F**). Overall, these results demonstrate that patients on active B cell-targeted therapies exhibited substantively diminished booster responses, as measured by induction of neutralizing antibodies. Furthermore, SARS-CoV2 mRNA vaccination using the ancestral spike protein was able to induce neutralizing antibodies against Omicron in many patients, although predominantly in patients without active therapy.

### Clinical correlates and monoclonal antibody therapy

In depth analysis of clinical parameters associated with the detection of antibodies is provided in **Table S1**. Likely due to insufficient patient numbers, we were unable to draw significant conclusions as to the impact of disease (CLL and non-Hodgkin’s lymphoma) or specific therapy, although these parameters are illustrated graphically (**Fig. S1**). There was also not a clear association between clinical parameters such as type of treatment, gender, age, days post-booster, white blood cell (WBC) counts at booster, and absolute lymphocyte counts (ALC) at booster. Graphical display of longitudinal analysis of all study patients plotted versus their booster timepoint illustrated the impact of active therapy on antibody levels (**Fig. S3**).

As an illustrative example of the impact of mAb treatment, we assessed additional post-booster samples taken from three patients who were initially seronegative after primary vaccination. These patients tested positive for infection with SARS-CoV2, and within 4 days, were treated with the Regeneron mAb cocktail (Casirivimab and Imdevimab). Full details are provided in **Figure S4**. In these patients, we detected remarkable antibody selectivity for ancestral RBD in comparison with Omicron RBD, consistent with recent reports demonstrating that the Regeneron mAb cocktail does not efficiently recognize Omicron RBD (1-3, 27). These results are consistent with detection of infused antibodies and lack of endogenous antibody responses despite booster and infection. Our results are noteworthy as many immunosuppressed patients are likely to be treated with mAb therapy which may be impactful on antibody correlative studies.

### B cell targeted therapy selectively impacts de novo versus pre-existing antibody levels

An important question is whether B cell-targeted therapies impair the maintenance of pre-existing antibody levels as well as de novo antibody generation. To assess this question, we measured relative levels of antibodies against (i) EBV (gp350), (ii) Influenza, H1N1, A/Brisbane/2018 (HA protein), and (iii) the common cold coronavirus OC43 (spike), in addition to the ancestral RBD (**Fig. 4A-D**). In contrast to antibodies against SARS-CoV2 RBD, those specific for EBV, influenza, and the common coronavirus were not significantly different in patients with active treatment versus no active treatment. Importantly, there was no correlation between lack of ancestral RBD antibodies and lack of antibodies to any of the other viral targets (**Fig. 4E-G**). More extensive analysis is provided in **Figure S5**, including results obtained for other viral targets, and patients subdivided into groups with different levels of vaccine response against the ancestral RBD protein. These results demonstrate pre-existing antibody levels against endemic viruses were not altered even in patients without detectable antibodies to ancestral RBD following SARS-CoV2 vaccination and booster.

**Figure 4:**
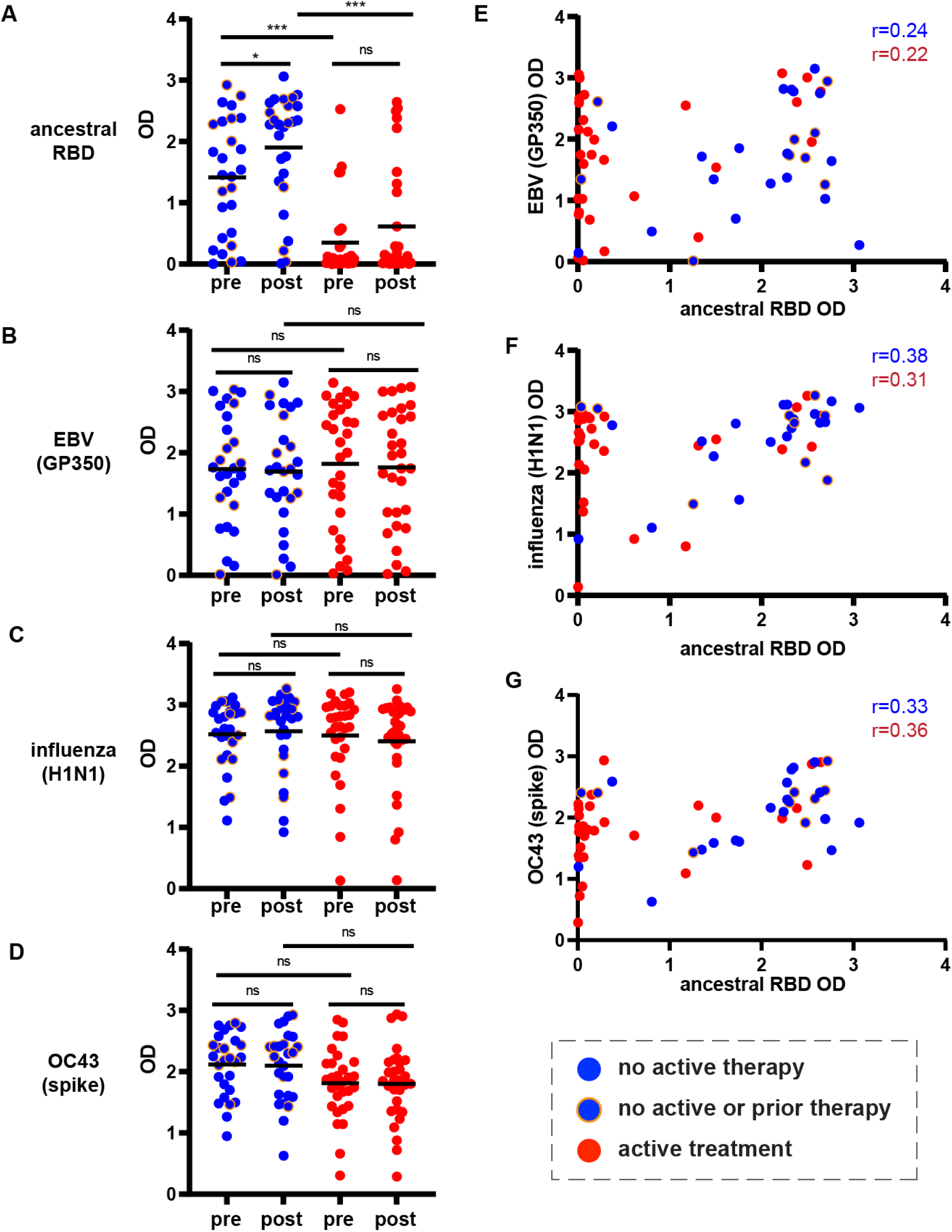
B cell-depleting therapy selectively impairs de novo antibody generation. (A) Shows antibody levels determined by ELISA against the ancestral RBD protein. Each patient is represented by a dot pre- and post-therapy. Patients without active therapy are indicated in blue while patients on active therapy are indicated in red. The orange circle around the blue dots indicates patients that are untreated. The arithmetic mean is shown with the bar. (B) As in ‘A’, except antibodies against the EBV GP350. (C) As in ‘A’ except antibodies against influenza H1N1, and (D) As in ‘A’ except antibodies against the OC43 common cold coronavirus spike protein. (E) Shows a dot plot of ancestral RBD versus EBV GP350 antibody binding taken from post-booster data. (F) As in ‘E’ except ancestral RBD versus influenza H1N1. (G) As in ‘E’ except ancestral RBD versus OC43 spike. For A-D, the Wilcoxon signed-rank test was used to compare OD values between pre- and post-booster while Mann-Whitney test was used to compare between different conditions (treated and non-treated), and the Bonferroni correction was applied. *, *** indicate p< 0.05, 0.001 For E-G, the Spearman’s rank correlation coefficient was calculated.

## Discussion

We report substantial heterogeneity in antibody responses following booster immunization in patients with B cell malignancies. While patients not on active cancer therapy show significant increases in antibody levels after booster immunization, many patients on active therapy remain seronegative despite primary immunization and booster. Importantly, among all seropositive patients, there was a strong correlation between antibodies against the ancestral strain and Omicron variant. These findings indicate that Omicron is not sufficiently antigenically distinct to escape all vaccine-mediated antibody immunity. However, as previously reported in healthy individuals and solid tumor patients (2-4, 11, 12, 15-17), in this study neutralizing antibody levels against Omicron were reduced in comparison with the ancestral virus. Altogether, our results build on previous findings showing that B cell-targeted therapies are associated with reduced antibody responses after primary vaccination (20, 22-24, 28, 29) and that a significant fraction of patients with B cell malignancies remain seronegative after booster immunization (12, 17, 23, 25, 30). Our findings contrast with the studies in solid tumor patients, where the majority of patients develop robust booster-mediated responses, although cytoreductive chemotherapies may impair responses in select patients (11, 18, 20-22, 31). While our current study has focused on antibody levels, antigen-specific B and T cells are likely also critical for protective immunity. Of note, memory B cell levels can increase while antibody levels decrease (32). In multiple sclerosis patients receiving anti-CD20 mAb, there were elevated SARS-CoV2-reactive CD8^+^ T cell responses (29), suggesting compensatory immune pathways. Recent findings also show T cell cross reactivity between the ancestral and Omicron variant (33, 34). Altogether, our results demonstrate that booster immunization may be beneficial to select patients with B cell malignancies, however, further studies will be needed to determine if post-booster seronegative patients may also benefit from their vaccination regimen.

A second important finding in our study was that regardless of active therapy or seronegative status, there were near normal levels of pre-existing antibodies against endemic viruses such as influenza and EBV. While many studies in humans in the pre-SARS-CoV2 era have shown that B cell-targeted therapies impair vaccine-mediated antibody responses (35-39), few studies have assessed pre-existing antibody levels. In mice and macaques, pre-existing antibodies, including those generated following vaccination and infection, were preserved after anti-CD20 mAb treatment (40, 41). In a small number of rheumatoid arthritis patients treated with anti-CD20 mAb, levels of antibodies against tetanus toxoid and pneumococcal capsular polysaccharides appeared to be maintained (42). Similar findings were also reported in multiple sclerosis patients treated with anti-CD20 mAb, although these findings were only reported as part of an abstract and in a digital repository (35, 43). Our results and these prior findings suggest B cell-targeted therapies have minimal impact on long-lived plasma cells (LLPCs), which are likely the source of basal and pre-existing antibody levels (44, 45). The ability of LLPCs to avoid anti-CD20 mAb and BTKi therapy may reflect lack of CD20 expression, tissue sequestration from drug, and signal-independent expression of antibodies. While our findings suggest that B cell-targeted therapies do not impair the maintenance of pre-existing antibody levels, future studies will be needed to assess protective immunity. However, our data suggest that vaccination prior to B cell-targeted therapy may be important to achieve long-term benefits. In support of this notion, two recent studies, with a relatively small number of patients, provide evidence that SARS-CoV2 vaccination prior to anti-CD20 mAb had superior induction of antibody responses compared to when the agents were administered in reverse order (46, 47). In summary, our findings highlight the discordance between de novo and pre-existing antibodies. To our knowledge, there have been no studies in humans showing that vaccine seronegative patients can achieve normal antibody levels against endemic viruses. Our findings provide a framework for further studies to understand antibody biology and to develop more effective vaccine strategies in patients with B cell-targeted therapies. Finally, our work suggests B cell malignancy patients on active therapy may be disproportionally vulnerable to emerging or new infections because of an inability to generate de novo antibody responses.

### Limitation of our study

An important limitation in our study is that while active therapy is associated with blunted antibody responses, we cannot formally determine whether reduced antibody responses are due to therapy or more advanced disease. Other limitations of our study include a relatively low patient number, the inherent heterogeneity in patients both in terms of disease and treatment, and the range in time between vaccination and blood draws. While we excluded patients with SARS-CoV2 therapeutic mAb treatment within 6 months of the post-booster blood draw, as our patient population is highly vulnerable, medical records may not have reflected mAb therapy performed at other sites. We also did not assess other immune correlates that may be impactful for protection including memory B cells, the presence of which may be more critical than antibody levels. Finally, while our data suggest that it may be important to vaccinate prior to initiating B cell targeted therapy, clinical studies will be necessary to further validate this possibility.

## Methods

### Human subjects

Serum samples were collected from cancer patients enrolled under approved IRB protocol (2021C004) as part of the SIIREN study (The COVID-19 Vaccine <u>S</u>tudy of Infections and Immune <u>RE</u>spo<u>N</u>se) at The Ohio State University Comprehensive Cancer Center. All cancer patients received two mRNA vaccine doses and 87 to 276 days later received an additional (third) mRNA booster dose. Cancer patient sera (n=57) was collected pre- and post-booster mRNA vaccine. Pre-booster samples were collected between 66 and 216 days post second mRNA vaccine, and post-booster samples were collected between 5 and 112 days after mRNA booster. All cancer patients had diagnoses of B cell malignancies including CLL (n=35) or non-Hodgkin’s Lymphoma (NHL) (n=22). Patients were separated into those not on active therapy (n=27) and on active therapy (n=30) based on B cell-targeted therapy being administered within 9 months of the primary vaccination. Two patients were included in the study who received mAb therapy within 3 months prior to primary vaccination. Clinical information was extracted from the internal electronic medical record database, including age, sex, race, cancer diagnosis and therapy status, COVID-19 mRNA vaccine information, clinical laboratory findings, and diagnosis/treatment of COVID-19 infection. All patient demographic and clinical data is further described in Table 1. One patient removed from the main cohort but reported in supplemental Figure 4 was excluded due to administration of COVID-19 monoclonal antibody shortly prior to post-booster sample collection (∼ 8 days).

### ELISAs

ELISAs were conducted as previously described (48). Briefly, the wells of 96-well plates (Costar) were coated with 50 µL of 2 µg/mL of target protein in 1x PBS (VWR) overnight at 4°C. Target proteins include SARS-CoV-2 proteins (Ancestral spike, Lakepharma; Ancestral RBD, Lakepharma; Omicron spike, Sinobiological; Omicron RBD, Sinobiological; EBV (GP350), Sinobiological; Influenza A, H1N1, Brisbane/2018 (HA protein), Sinobiological; Influenza A, H3N2,Comabodia/2020 (HA protein), Sinobiological; OC43 (spike), Sinobiological; HKU1 (spike), Sinobiological; NL63 (spike), Sinobiological; and 229E (spike), Sinobiological). The next day, the protein coating solution was removed and the plates were washed 3x with PBS with 0.1% Tween 20 (Millipore Sigma) using the Elx405 automated plate washer (Biotek). Each well was filled with 200 µL of a blocking PBS-T solution with 3% dry powder milk (Fisher Scientific) and incubated at room temperature for 2 hours. Then, the blocking solution was removed, and the plates were blotted on a paper towel to remove residual blocking buffer. Serum dilutions were prepared the same day using a PBS-T and 1% milk powder solution and dispensed into the plate at 100 µL per well and incubated at room temperature for 2 hours. (Prior to use, serum was heat inactivated by incubation at 56°C for 1 hour.) Next, the plates were washed 3x with 0.01% PBS-T and 50 µL of an HRP-conjugated Anti-Human IgG, IgA, or IgM (Goat Anti-Human IgG/IgA/IgM Fc crossed-absorbed secondary antibody HRP, Invitrogen, used at 1:3000 dilution) antibody solution in 1% milk powder and PBS-T solution was added to each well and incubated in the dark at room temperature for 1 hour. After incubation, the solution was removed and the plates were washed 3x more with 0.1% PBS-T. Immediately after adding 1 mL of 10x stable peroxide substrate (Thermofisher) to a solution of one o-phenylenediamine dihydrochloride tablet (Thermofischer) in 9 mL of diH20, 100 µL was dispensed into each well. This solution developed in the plates for 10 minutes before adding 50 µL of 2.5 M sulfuric acid solution (Fischer Scientific) to each well to stop the reaction. Once stopped, the optical density (OD) values for the plates were read at 490 nm using the SpectraMax iD5 Multi-Mode Microplate Reader (Molecular Devices). ELISAs for the SARS-CoV-2 target proteins (spike, RBD, Omicron spike, Omicron RBD) were performed in one run per target protein with three dilutions (1:200, 1:800, and 1:3200). Controls were assigned to every plate to account for inter-plate variability. The diluents average OD was subtracted to obtain the final OD values of the samples. Area under the curve (AUC) was calculated using Graphpad Prism 9. ELISAs for other targets was performed at 1:200 and each target was assessed in two independent assays with similar results. For determination of detectable antibody levels, the cut-off was determined as 3 standard deviations above the mean of the negative control samples.

### Neutralization assay

Pseudotyped virus neutralization assays were performed as previously described (20, 26). Briefly, patient serum was 4-fold serially diluted and incubated with equivalent amounts of infectious D614G or Omicron (B.1.1.529) pseudotyped virus (final dilutions of 1:40, 1:160, 1:640, 1:2560, 1:10240, and no serum control). Following 1hr incubation at 37°C, virus was used to infect HEK293T-ACE2 cells. Gaussia luciferase activity was assayed 48hr and 72hr after infection by combining 20 μL of cell culture media with 20 μL of Gaussia luciferase substrate (0.1M Tris pH 7.4, 0.3M sodium ascorbate, 10 μM coelenterazine). Luminescence was measured by a BioTek Cytation5 plate reader. Neutralization curves were plotted in GraphPad Prism 5 using least-squares fit non-linear regression to determine neutralizing titers 50%.

### Constructs

The construct used for the production of lentiviral pseudotypes was pNL4-3-ΔEnv-inGluc (49), which was originally obtained from David Derse’s lab at NIH (National Cancer Institute, Frederick, Maryland, USA) and Marc Johnson’s lab at the University of Missouri (Columbia, Missouri, USA). This construct is based on a ΔEnv pNL4.3 HIV-1 vector and contains an anti-sense Gaussia luciferase (Gluc) gene with a sense intron. Gluc is secreted in mammalian cell culture, and the intron and anti-sense orientation of the Gluc gene prevents the production of Gluc in the virus producer cells. The codon-optimized D614G and Omicron (B.1.1.529) SARS-CoV-2 S constructs were synthesized by GenScript (Piscataway, NJ) and subsequently cloned into a pcDNA3.1 vector by restriction enzyme cloning with Kpn I and BamH I.

### Cell Lines and Maintenance

HEK293T cells (ATCC CRL-11268, CVCL_1926) and HEK293T-ACE2 cells (BEI, NR-52511) were maintained in DMEM (Gibco, 11965-092) supplemented with 10% FBS (Sigma, F1051) and 1% penicillin/streptomycin (HyClone, SV30010). Both cell lines were maintained at 37°C, and 5% CO2 in tissue culture treated, sterile 10cm dishes (FisherScientific, FB012924).

### Pseudotyped Virus Production

Pseudotyped virus was produced as previously described (20, 26). Pseudotyped lentivirus was produced by co-transfection of HEK293T cells with pNL4-3-ΔEnv-inGluc and D614G or Omicron spike expression constructs in a 2:1 ratio by polyethylenimine (PEI) transfection. Pseudotyped virus was collected 24, 48, and 72hrs after transfection, pooled, aliquoted, and stored at -80°C. Relative pseudotyped virus infectivity was assessed by infection of HEK293T-ACE2 cells. Media was assayed for Gaussia luciferase activity by combining 20 μL of cell culture media with 20 μL of Gaussia luciferase substrate (0.1M Tris pH 7.4, 0.3M sodium ascorbate, 10 μM coelenterazine). Luminescence was measured by a BioTek Cytation5 plate reader.

### Pseudotyped Virus Neutralization Assay

Pseudotyped virus neutralization assays were performed as previously described (20, 26). Patient serum was 4-fold serially diluted and incubated with equivalent amounts of infectious D614G or Omicron (B.1.1.529) pseudotyped virus (final dilutions of 1:40, 1:160, 1:640, 1:2560, 1:10240, and no serum control). Following 1hr incubation at 37°C, virus was used to infect HEK293T-ACE2 cells. Gaussia luciferase activity was assayed 48hr and 72hr after infection by combining 20 μL of cell culture media with 20 μL of Gaussia luciferase substrate. Luminescence was measured by a BioTek Cytation5 plate reader. Neutralization curves were plotted in GraphPad Prism 5 using least-squares fit non-linear regression to determine neutralizing titers 50%.

### Statistical Analysis

GraphPad Prism 9 and R 4.1.1 were used for statistical analyses. Wilcoxon signed-rank test was used to compare paired values and Mann-Whitney test was used to compare unpaired values. Multiple testing was adjusted using the Bonferroni correction. ANOVA was used for multiple group comparisons. For correlation studies, Spearman’s rank correlation was used. Fisher’s exact test was used for association analysis between two categorical variables. The significance level of 0.05 was used to determine significance; * indicates p < 0.05, ** p < 0.01, *** p < 0.001 and **** p < 0.0001. Statistical details for each graph or table are provided in the figure legends.

## Data Availability

All data produced in the present study are available upon reasonable request to the authors

## Acknowledgments

We are grateful for the patients who participated in the OSU SIIREN study. We are also grateful to the following people who helped support clinical efforts related to the trial: Kambra Crist, Kayla Swetel, Caroline Gault, Taylor Surplus, Alek Erdel, Abdelhalim Belkheyar, Benjamin Veneman, David Cohn, Kristin Thatcher, Michele Peddicord, Carolyn McClerking, Daniel Weng, Amarnath Singh, Mallory Dietrich, and Josh Van Le. We are also grateful for the OSUMC/James clinical staff members and members of the OSUWMC Clinical Research Center who dedicated their time to assist sample collection. We also thank members of the Liu lab, including Yi-Min Zheng, Panke Qu, and Julia Faraone, for sharing reagents and discussion. Additional gratitude is extended toward Kelsi Reynolds, Mohamed Yusuf, Robert Davenport, and Taylor Chatlos from the Immune Monitoring and Discovery Platform of the Pelotonia Institute for Immuno-Oncology for assistance in sample collection and discussion. We thank the Recruitment, Intervention and Survey Shared Resource at The Ohio State University Comprehensive Cancer Center for data management. We thank Serge Maalouf for helpful discussions. Finally, we would like to thank the NIH AIDS Reagent Program and BEI Resources for supplying important reagents that made this work possible.

Research reported in this publication was supported by The Ohio State University Comprehensive Cancer Center and the National Institutes of Health under grant number P30 CA016058. S-LL was supported by an NIH grant R01 AI150473, by the National Cancer Institute of the NIH under award no. U54CA260582, and by a fund provided by an anonymous private donor to OSU. JPE was supported by Glenn Barber Fellowship from the Ohio State University College of Veterinary Medicine. MPR was supported by NIH grant R01CA222817.

