## Supplementary material for "Suppression of de novo antibody responses against SARS-CoV2 and the Omicron variant after mRNA vaccination and booster in patients with B cell malignancies undergoing active treatment, but maintenance of pre-existing antibody levels against endemic viruses": sup figures and table

### Figure S1

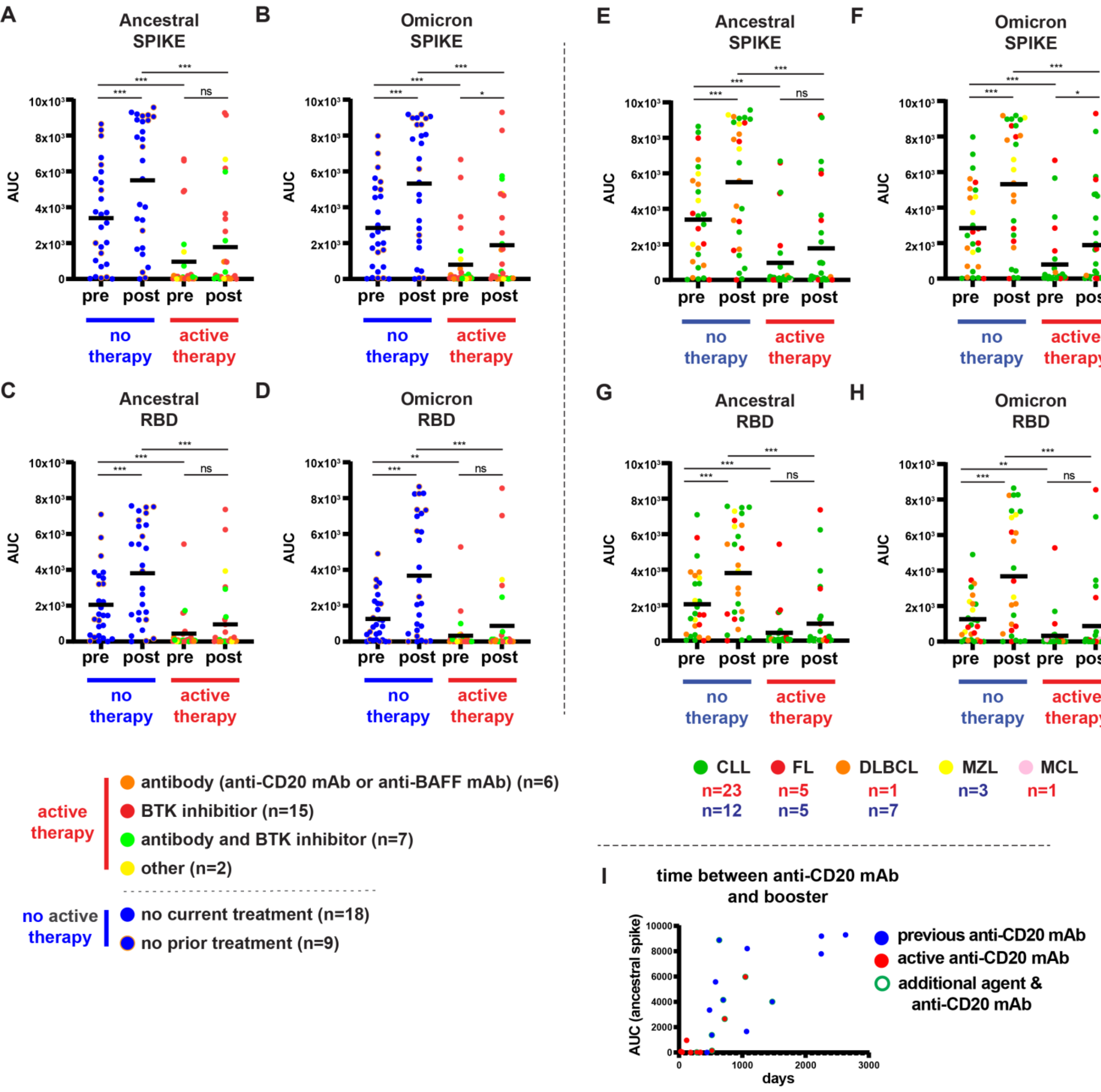

A
Ancestral SPIKE
B
Omicron SPIKE
C
Ancestral RBD
D
Omicron RBD
E
Ancestral SPIKE
F
Omicron SPIKE
G
Ancestral RBD
H
Omicron RBD
AUC
pre
post
no therapy
active therapy
active therapy
antibody (anti-CD20 mAb or anti-BAFF mAb) (n=6)
BTK inhibitior (n=15)
antibody and BTK inhibitor (n=7)
other (n=2)
no active therapy
no current treatment (n=18)
no prior treatment (n=9)
CLL
FL
DLBCL
MZL
MCL
n=23
n=5
n=1
n=3
n=1
n=12
n=5
n=7
I
time between anti-CD20 mAb and booster
AUC (ancestral spike)
days
previous anti-CD20 mAb
active anti-CD20 mAb
additional agent & anti-CD20 mAb

**Figure S1: ELISA data showing antibody levels against the ancestral and Omicron proteins with patients color coded by treatment and time.** (A-D) Patients on active treatment are color coded based on their therapy including monoclonal antibodies against CD20 or BAFF (orange), BTK inhibitors (red), the combination of the latter (green), and other therapies (yellow) which includes anti-metabolite (lenalidomide) and BCL-2 inhibitor monotherapy. For patients not on active therapy (blue), untreated patients have an orange circle. The graphs depict antibodies against (A) ancestral spike, (B) Omicron spike, (C) ancestral RBD, and (D) the Omicron RBD before and after booster vaccination in patients with or without active therapy. Each point represents a patient, and the bar represents the arithmetic mean. The Wilcoxon signed-rank test was used to compare AUC values between pre- and post-booster while Mann-Whitney test was used to compare between different conditions (treated vs non-treated), and the Bonferroni correction was applied. *, **, *** indicate $p < 0.05, 0.01, 0.001$. (E-H) Same as ‘A-D’, except patients are color coded by lymphoma type including CLL and Non-Hodgkin lymphoma subtypes (FL (Follicular lymphoma), DLBCL (Diffuse large B-cell lympoma), MZL (Marginal zone lymphoma), and MCL (Mantle cell lymphoma)). The red numbers indicate the number of patients for each lymphoma type in the active therapy group while the blue numbers indicate the number of patients for each lymphoma type in the not active therapy group . (I) All patients receiving anti-CD20 mAb therapy were graphed with ancestral spike antibody levels (AUC) after booster administration versus time between last anti-CD20 mAb administration and booster.

### Figure S2

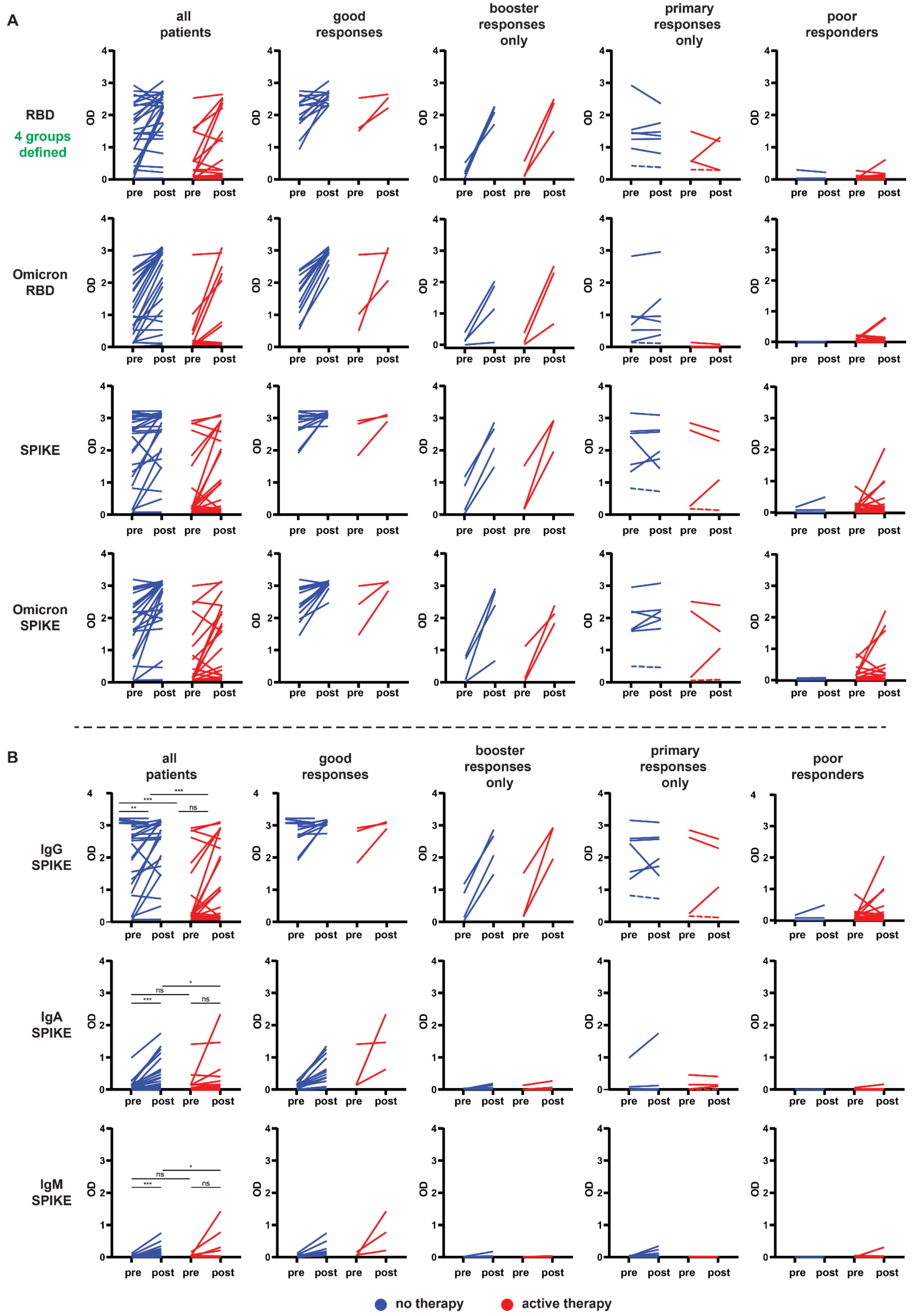

A
all patients
good responses
booster responses only
primary responses only
poor responders
RBD
4 groups defined
Omicron RBD
SPIKE
Omicron SPIKE
OD
pre
post
B
IgG SPIKE
IgA SPIKE
IgM SPIKE
***
**
ns
*
no therapy
active therapy

**Figure S2: Patients grouped for comparison by vaccine response as measured by antibodies against the ancestral RBD.** (A) These graphs depict (from left to right) all patients, patients with a good response to primary and booster vaccination, patients with a good response to the booster, patients with a good primary response, and patients with a relatively poor response to vaccination. Groups were defined based on the ancestral RBD response. Antibody levels were estimated by OD measurement at a 1:200 serum dilution. Each line represents one patient and a paired pre- and post- booster timepoint. The dashed line in the "primary response" group represents the same patients in each condition for this and subsequent graphs. Dashed lines represent the patients with the lowest IgG OD readings in the primary response only group. Please refer to Figure S1 for statistics relevant to these conditions. (B) Patients were grouped as outlined in 'A'. Shown are detection of IgG, IgA, and IgM antibodies reactive against the ancestral spike protein. Wilcoxon signed-rank test was used to compare OD values between pre- and post-booster while Mann-Whitney test was used to compare between different conditions (treated and non-treated), and the Bonferroni correction was applied. *, **, *** indicate $p < 0.05, 0.01, 0.001$

### Figure S3

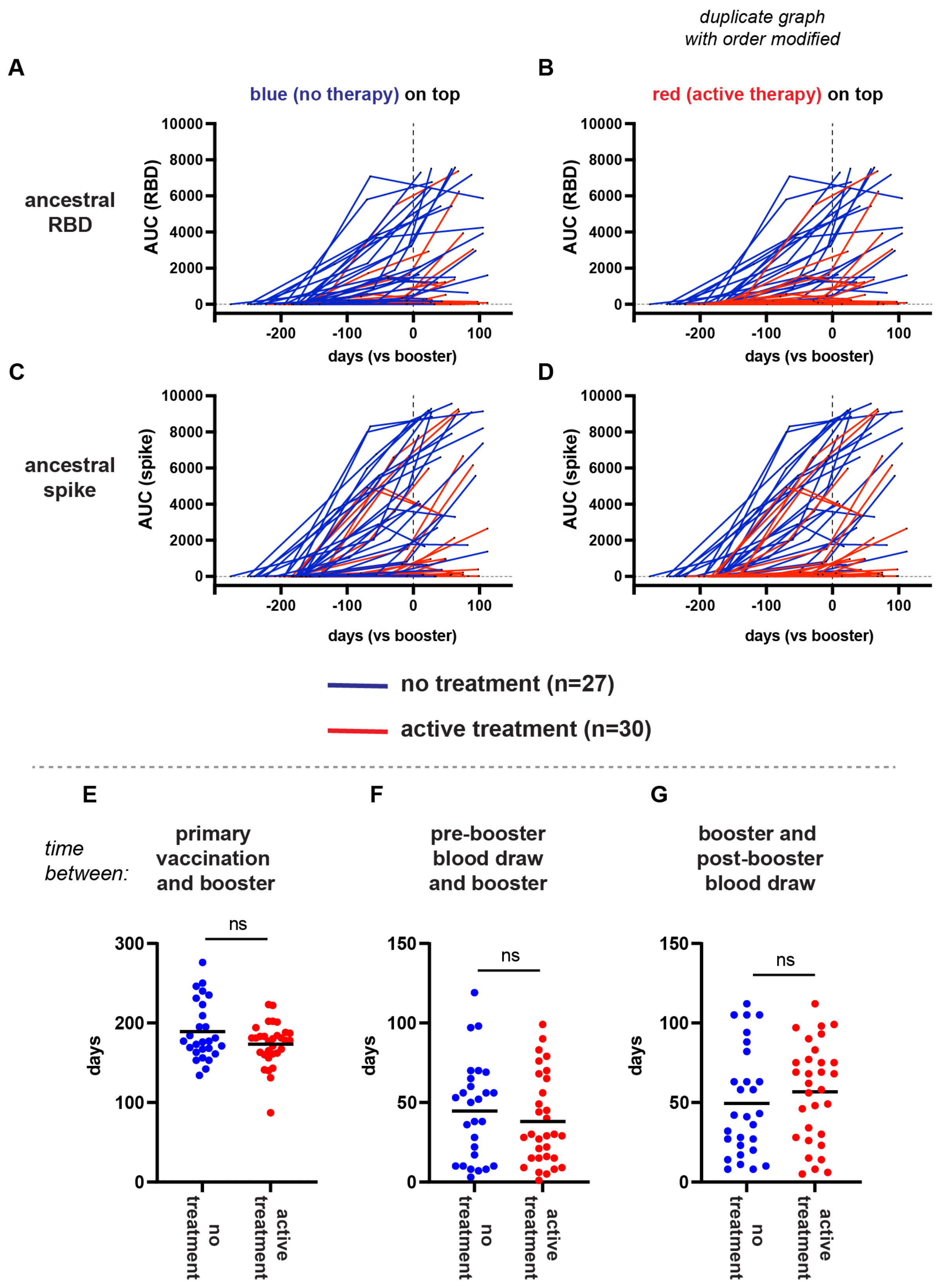
duplicate graph
with order modified
A
B
blue (no therapy) on top
red (active therapy) on top
ancestral
RBD
AUC (RBD)
10000
8000
6000
4000
2000
0
-200
-100
0
100
days (vs booster)
C
D
ancestral
spike
AUC (spike)
no treatment (n=27)
active treatment (n=30)
E
F
G
time
between:
primary
vaccination
and booster
pre-booster
blood draw
and booster
booster and
post-booster
blood draw
ns
days
300
200
100
150
50
no
treatment
active
treatment

**Figure S3: Graphical depiction of ancestral RBD and spike antibody responses in relationship to time.** (A) Shows antibody responses (AUC) graphed in relationship to days before or after the booster. Each line represents one patients and includes (i) primary vaccination defined as 0, (ii) the pre-booster timepoint, and (iii) the post-booster time point. Some patients had additional timepoints that were also included although extra timepoints from Figure S4 were not included. (B) Shows identical data to 'A' except the red lines are on top instead of the blue lines. (C & D) Identical to 'A & B' except using ancestral spike as the target. (E) Shows time between the primary vaccination (2nd timepoint) and the booster. (F) Shows the time between pre-booster timepoint and the booster. (G) Shows the time between the booster and the post-booster blood draw. For E-G, each dot represents a patient and the bar represents the arithmetic mean. The Mann-Whitney test was used to compare between active and no treatment in E,F, & G.

### Table S1

#### Table S1. Correlation of clinical parameters with NT50 antibody levels

**A**

| PRE-BOOSTER | Ancestral | | Omicron | |
|---|---|---|---|---|
| Parameter | No Treatment | Treatment | No Treatment | Treatment |
| Age | −0.057 (0.778) | −0.227 (0.228) | 0.194 (0.331) | 0.088 (0.643) |
| Days after booster | −0.04 (0.844) | 0.001 (0.996) | 0.129 (0.521) | 0.017 (0.929) |
| Log of WBC | 0.237 (0.234) | 0.059 (0.758) | 0.309 (0.117) | -0.299 (0.108) |
| Log of ALC | 0.228 (0.252) | 0.023 (0.904) | 0.29 (0.142) | -0.288 (0.123) |

| POST-BOOSTER | Ancestral | | Omicron | |
|---|---|---|---|---|
| Parameter | No Treatment | Treatment | No Treatment | Treatment |
| Age | -0.053 (0.792) | -0.141 (0.457) | -0.089 (0.658) | 0.059 (0.758) |
| Days after booster | -0.023 (0.908) | 0.238 (0.206) | -0.001 (0.997) | 0.237 (0.208) |
| Log of WBC | 0.241 (0.227) | 0.008 (0.965) | 0.276 (0.164) | -0.057 (0.766) |
| Log of ALC | 0.23 (0.249) | 0.029 (0.881) | 0.282 (0.154) | -0.103 (0.589) |

| PRE-POST DIFF | Ancestral | | Omicron | |
|---|---|---|---|---|
| Parameter | No Treatment | Treatment | No Treatment | Treatment |
| Age | -0.022 (0.914) | 0.021 (0.914) | -0.177 (0.378) | -0.009 (0.963) |
| Days after booster | 0.008 (0.97) | 0.277 (0.138) | -0.057 (0.779) | 0.224 (0.235) |
| Log of WBC | 0.123 (0.542) | -0.038 (0.841) | 0.154 (0.443) | 0.172 (0.362) |
| Log of ALC | 0.115 (0.568) | 0.015 (0.939) | 0.169 (0.401) | 0.118 (0.535) |

Values are listed as: Correlation (P value)

---

**B**

| PRE-BOOSTER | Ancestral | | Omicron | |
|---|---|---|---|---|
| Parameter | No Treatment | Treatment | No Treatment | Treatment |
| Sex | 0.941 | 0.659 | 0.981 | 0.174 |
| Vaccine Type | 0.626 | 0.209 | 0.277 | 0.680 |

| POST-BOOSTER | Ancestral | | Omicron | |
|---|---|---|---|---|
| Parameter | No Treatment | Treatment | No Treatment | Treatment |
| Sex | 0.716 | 0.659 | 0.790 | 0.980 |
| Vaccine Type | 0.323 | 0.615 | 0.486 | 0.805 |

| PRE-POST DIFF | Ancestral | | Omicron | |
|---|---|---|---|---|
| Parameter | No Treatment | Treatment | No Treatment | Treatment |
| Sex | 0.422 | 0.527 | 0.981 | 0.082 |
| Vaccine Type | 0.373 | 0.805 | 0.516 | 0.563 |

Values are listed as: P value

**Table S1: Correlation of clinical parameters with $NT_{50}$ antibody levels** Association of clinical parameters and $NT_{50}$ values at the pre-booster timepoint, post-booster timepoint), or the difference between pre- and post- booster. (A) Shows correlation values with P-values in parenthesis, and (B) shows P-values using the Mann-Whitney test . $NT_{50}$ values were first log10-transformed and then difference of log10($NT_{50}$) values between post- and pre-booster were calculated. Mann-Whitney was used for clinical variables with two categories. Pearson correlation analysis was used for continuous clinical variables. When multiple pairs are simultaneously considered, multiple testing was adjusted using the Bonferroni correction.

### Figure S4

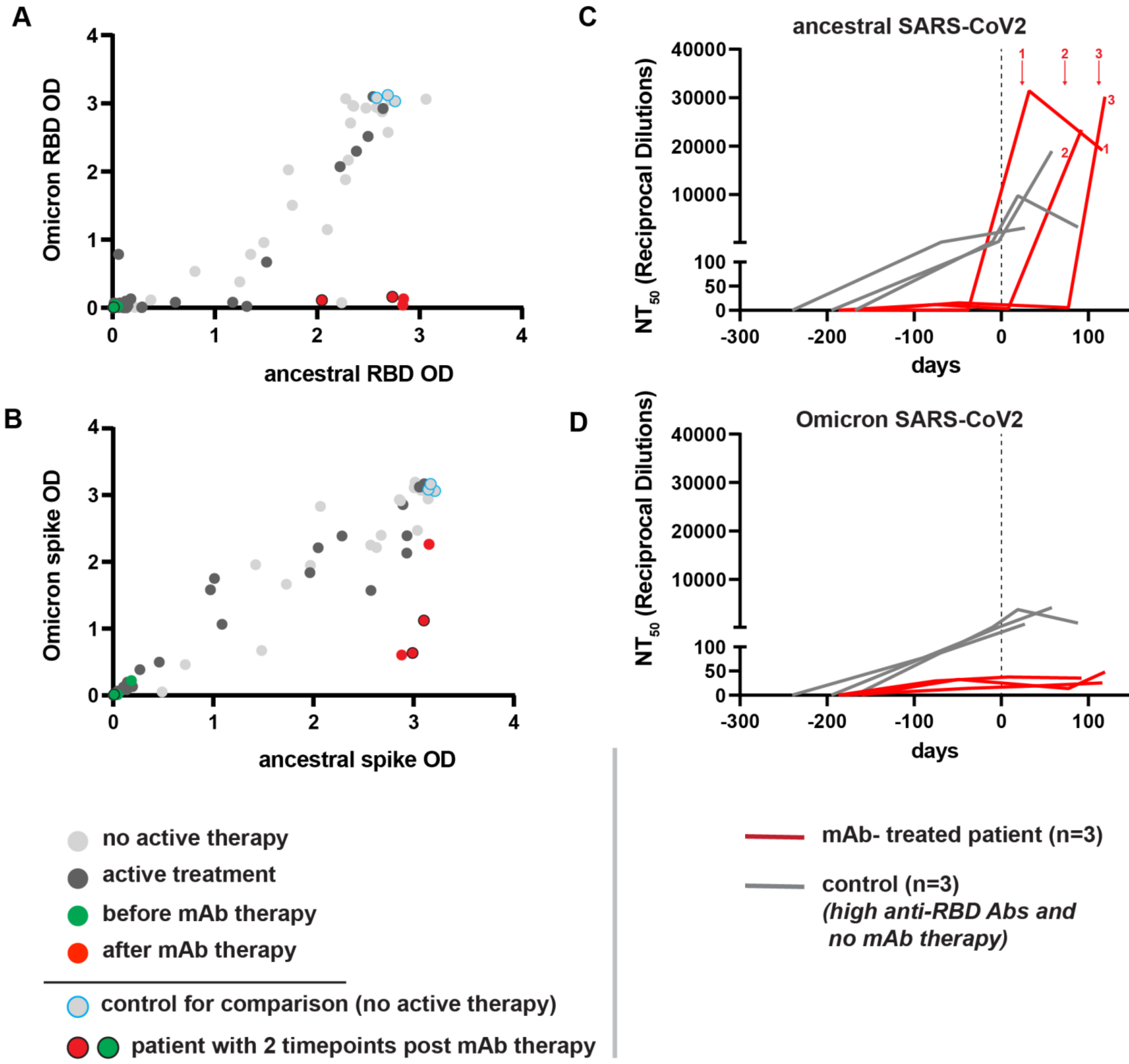
A
Omicron RBD OD
ancestral RBD OD
0
1
2
3
4
B
Omicron spike OD
ancestral spike OD
C
ancestral SARS-CoV2
NT50 (Reciprocal Dilutions)
40000
30000
20000
10000
100
50
-300
-200
-100
days
D
Omicron SARS-CoV2
no active therapy
active treatment
before mAb therapy
after mAb therapy
control for comparison (no active therapy)
patient with 2 timepoints post mAb therapy
mAb- treated patient (n=3)
control (n=3)
(high anti-RBD Abs and
no mAb therapy)

**Figure S4: Antibody responses in patients receiving mAb therapy.** (A) Dot plot shows the post-booster patient timepoints graphed by ancestral RBD versus Omicron RBD as determined by ELISA. Additional timepoints (red and green) have been added onto the dot plot to (i) highlight post-booster and pre mAb therapy timepoints for selected patients, (ii) post-booster and post mAb therapy timepoints for selected patients, and (iii) three timepoints from an additional patient with 1 analysis pre mAb therapy and 2 analyses post mAb therapy. (B) Similar to 'A' except showing ancestral spike versus Omicron spike ELISA values. (C) Shows neutralization assay and NT50 values for neutralization against the ancestral SARS-CoV2 virus. The red arrows indicate when mAb therapy was administered for the 3 patients. (D) as in 'C' except the Omicron variant of SARS-CoV2.

### Figure S5

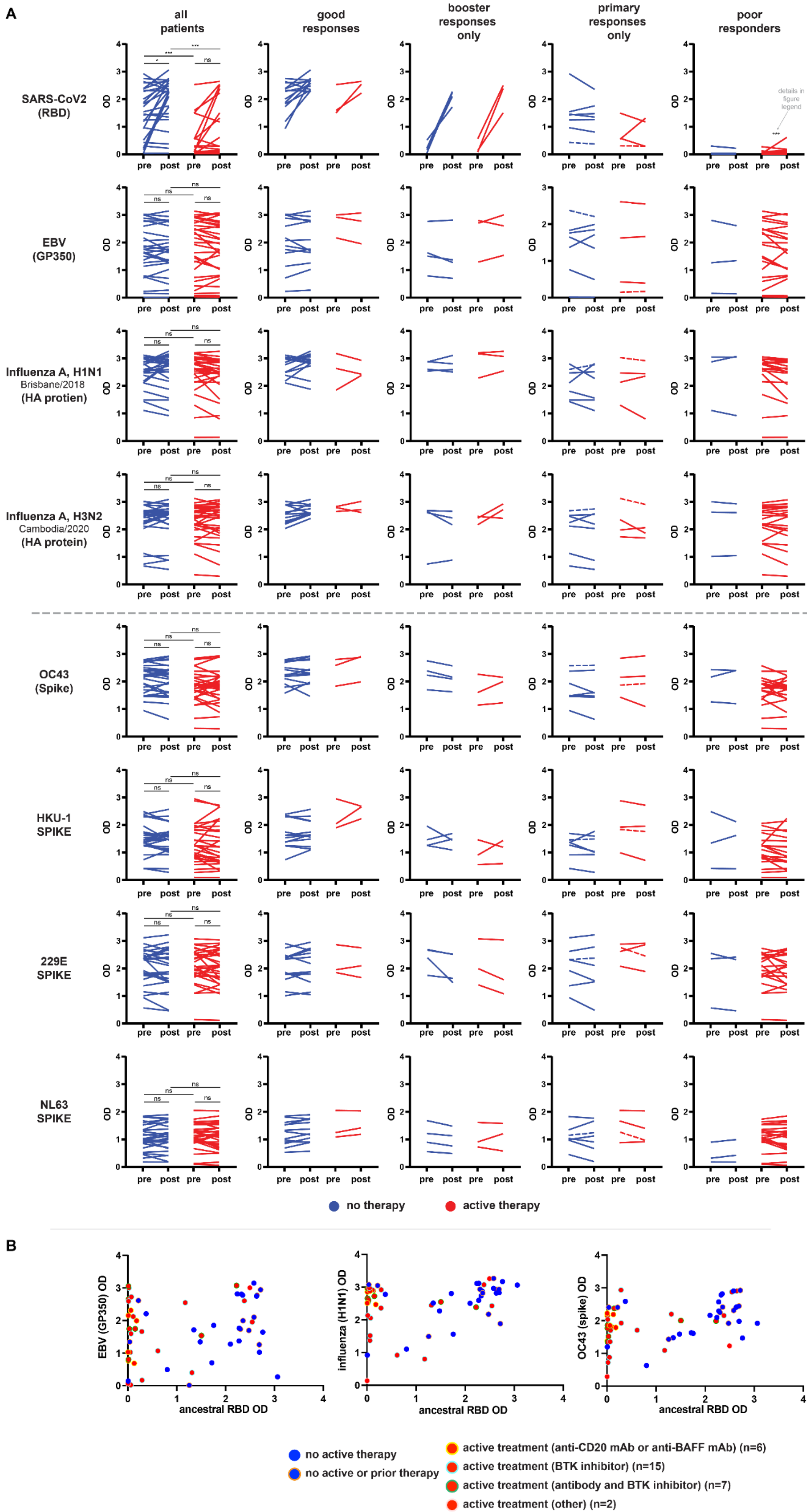
A
all patients
good responses
booster responses only
primary responses only
poor responders
SARS-CoV2 (RBD)
EBV (GP350)
Influenza A, H1N1
Brisbane/2018
(HA protien)
Influenza A, H3N2
Cambodia/2020
(HA protein)
OC43 (Spike)
HKU-1 SPIKE
229E SPIKE
NL63 SPIKE
OD
pre
post
ns
details in figure legend
no therapy
active therapy
B
EBV (GP350) OD
influenza (H1N1) OD
OC43 (spike) OD
ancestral RBD OD
no active therapy
no active or prior therapy
active treatment (anti-CD20 mAb or anti-BAFF mAb) (n=6)
active treatment (BTK inhibitor) (n=15)
active treatment (antibody and BTK inhibitor) (n=7)
active treatment (other) (n=2)

**Figure S5: B cell-depleting therapy selectively impairs de novo antibody generation.** (A) Patients were grouped as outlined in Figure S2 by vaccine response to the ancestral RBD. Shown is ELISA data for each of the viral targets outlined in the left column, including (top to bottom) ancestral RBD, EBV (GP350), H1N1 influenza (Brisbane, 2019-2020), H3N2 influenza (Cambodia, 2021-2022), OC43 spike, HKU-1 spike, 229E spike, and NL63 spike. Each line represents one patient connecting a pre- and post- booster timepoint. Patients not receiving therapy (blue) versus those receiving therapy (red) are color coded. The 2 patients with the dashed lines (both in the primary response group) are similarly denoted across conditions for illustrative purposes. Antibody levels are estimated by optical density (OD). In the “all patients” column, the Wilcoxon signed-rank test was used to compare OD values between pre- and post-booster while Mann-Whitney test was used to compare between different conditions (treated and non-treated). Bonferroni correction was applied. *, *** indicate $p < 0.05$, 0.001 In the poor responder's column, we also compared post-booster OD values of the WT RBD in the treated group (row 1, far right, red) with each of the OD values of the viruses in the treated group (rows 2-7, far right red) using the Wilcoxon signed-rank test. In all cases, this difference was significant (***, $p<0.001$) between row 1 and each of the other rows. (B) Identical to figure 4E, 4F, & 4G, but with treatment information in the active group color coded.
